# A Prospective Observational Study to Assess the Impact of Pharmacogenetics on Outcomes in Vascular Surgery (PROSPER)

**DOI:** 10.1101/2024.05.24.24307885

**Authors:** Kerry A Burke, Selman Mirza, Stuart J Wright, Nicholas S Greaves, William G Newman, John H McDermott

**Affiliations:** Manchester Centre for Genomic Medicine, St Mary’s Hospital, Manchester University NHS Foundation Trust, Oxford Road, Manchester, M13 9WL; Division of Evolution, Infection and Genomics, School of Biological Sciences, University of Manchester, Manchester M13 9PT; Manchester Vascular Centre, Manchester Royal Infirmary, Manchester University NHS Foundation Trust, Oxford Road, Manchester, M13 9WL; Centre for Biostatistics, School of Health Sciences, The University of Manchester, M13 9PL; Manchester Centre for Health Economics, The University of Manchester, Manchester, M13 9PL

**Keywords:** Pharmacogenetics, vascular surgery, clopidogrel, chronic limb threatening ischaemia

## Abstract

**Introduction:** Patients with chronic limb threatening ischaemia (CLTI) are often prescribed clopidogrel in order to reduce their risk of major adverse limb and cardiovascular events. Clopidogrel is metabolised by the CYP2C19 enzyme, and genetics variations in *CYP2C19* are common. These variants can influence an individual’s ability to metabolise clopidogrel to its active metabolite. This work aims to establish the relationship between patient genotype and outcomes after revascularisation in patients with CLTI who are prescribed clopidogrel. It will consider whether pharmacogenetics can be used to ensure patients are prescribed effective medications to optimise their outcomes.

**Methods and analysis:** This is a prospective observational cross-sectional study of patients undergoing lower limb surgical, endovascular or hybrid revascularisation for CLTI. Patients taking clopidogrel post-procedure, as well as those prescribed a non-clopidogrel based medication regimen, will be recruited prior to or shortly after revascularisation. Patients will undergo *CYP2C19* genotyping and will be followed-up using online records.

**Ethics and dissemination:** Manchester University Research Ethics Committee approval as obtained was part of the Implementing Pharmacogenetics to Improve Prescribing (IPTIP) trial process (IRAS 305751). The results of the study will be published in a peer-review journal and presented at international conferences.

**Registration:** This work is a sub-protocol for the IPTIP study which is registered as ISRCTN14050335.

## Introduction

Patients with chronic limb threatening ischaemia (CLTI) have significant lower extremity arterial disease and are known to be at high risk of major adverse limb events (MALE), including loss of vessel patency, need for surgical intervention and major amputation^1^. They are also at a significantly increased risk of major cardiovascular events (MACE), including myocardial infarction, cerebrovascular events and death^1^. Clopidogrel is an anti-platelet agent which is widely used in order to reduce the risk of both MALE and MACE in patients with CLTI^2,3^. It is a thienopyridine pro-drug which is metabolised by the CYP2C19 enzyme in the liver. Genetics variations in *CYP2C19* are common and can influence an individual’s ability to metabolise clopidogrel to its active metabolite^4^.

Research studies in both cardiac and stroke medicine have demonstrated significantly worse outcomes in patients treated with clopidogrel who are poor metabolisers of CYP2C19, and major guidelines have advocated a role of genetic testing in these specialties^5–7^. Research into the impact of *CYP2C19* alleles in vascular surgery is much more limited, but does suggest an association between poor metabolisers of CYP2C19 and adverse outcomes in patients taking clopidogrel^8^.

This protocol describes a prospective observational cross-sectional study which aims to establish the relationship between patient genotype and outcomes after revascularisation in patients with CLTI who are prescribed clopidogrel. It will consider whether pharmacogenetics can be used to ensure patients are prescribed effective medications to optimise their outcomes.

## Methods and analysis

This is a prospective observational cross-sectional study involving inpatients and outpatients at Manchester University NHS Foundation Trust (MFT), to assess whether genotype is associated with clinical outcome following revascularisation for patients with CLTI who are prescribed clopidogrel. A comparator group who are prescribed non-clopidogrel based medication regimens will also be recruited. This work is a sub-protocol for the Implementing Pharmacogenetics to Improve Prescribing trial (IPTIP) study^9^.

### Primary outcomes

1. Amputation-free survival at 1 year
2. Composite of Major Adverse Limb Events (MALE) in the index limb [amputation above the ankle, major limb re-intervention (graft revision, thrombectomy or thrombolysis), loss of graft/vessel patency] and death from any cause at 1 year

### Secondary outcomes

1. Minor re-intervention (angioplasty, stent)
2. MALE events at any time [amputation above the ankle, major limb re-intervention (graft revision, thrombectomy or thrombolysis), loss of graft/vessel patency]
3. Total re-interventions
4. Death within 30 days
5. Major adverse cardiovascular events (MACE) events at any time (myocardial infarction, cerebrovascular event or all-cause death)
6. Rate of systemic or gastro-intestinal bleed

### Inclusion criteria

- Patients awaiting revascularisation (either surgical, endovascular or hybrid) for CLTI or acute-on-chronic limb ischaemia, OR who have had a revascularisation procedure in the past 3 months
- No previous revascularisation in the study leg 3 months prior to this intervention (excluding diagnostic angiograms)
- Patients over the age of 18 years who are able to consent for themselves

### Exclusion criteria

- Patients receiving long term, full-dose anticoagulation post-procedure (does not include low-dose rivaroxaban)
- Acute limb ischaemia, aneurysmal disease, vasculitis, Buerger’s disease
- Patients being managed conservatively without revascularisation
- Patients who are pregnant, breastfeeding, on chemotherapy or radiotherapy
- Patients with less than six months life expectancy

Eligible participants will be provided with a patient information leaflet and will sign a consent form if they wish to enrol. A trained member of the research team will then take either a single blood sample or sputum sample, at the preference of the patient. Baseline demographic details will be collected during this time. Beyond this, participants will not be asked to do anything further for the study or to attend any future study visits. The blood or sputum sample will then be transported to the Manchester Centre for Genomic Medicine via existing and secure clinical pathways for internal specimen transport. DNA will be extracted and quantified by the North West Genomic Laboratory Hub (NW-GLH), a National Health Service (NHS) ISO15189 accredited laboratory. DNA samples will be labelled with the Study ID and stored at -20°C until genotyping. Genotyping will be undertaken using the AgenaTM iPLEX PGx 74 assay which can determine *CYP2C19* genotype status. All participants will have details about their past medical history, surgical history and prescribed medications collected from the Greater Manchester Care Record (GMCR). This information and the genetic data will then be linked in a secure database. This data will be pseudo-anonymised, meaning that the researcher analysing the data will not know whose data they are looking at.

The past medical history, surgical history and prescribed medication of participants will be recorded throughout the study period using the GMCR, NHS, MFT and other online records. The total follow-up period will be two years. Once the data collection period is complete, participant genotype will then be added to the final dataset within the secure research environment, linked by the Study ID. The final pseudonymised dataset will include the Study ID, patient demographics, past medical and surgical history, prescription record, patient genotype and metabolizer status.

Participants can withdraw consent at any time without giving any reason without their care or legal rights being affected, as participation in the research is voluntary. Patients who withdraw their consent to be part of the study will have their data removed from the final analysis pipeline and their DNA sample will be destroyed.

### Sample size

The desired sample size has been calculated assuming an average primary outcome rate of 0.35 at 1 year, a hazard ratio for *CYP2C19* loss of function (LoF) allele carriers for the primary endpoint of 2.0 and prevalence of LoF allele carriers of 26.2%^10,11^. Based on a survival analysis power calculation, using a two-tailed α of 0.05 and a β of 0.1, 114 events would be needed to identify an effect. Taking into consideration the baseline event rate, the study would need to recruit 326 participants receiving a clopidogrel-based medication regimen. Assuming 75% of all admissions receive a clopidogrel based antiplatelet therapy with 10% missing data, 483 participants would need to be recruited to the study in total.

### Statistical analysis

The whole cohort will be split into two groups for analysis. Group 1 involves patients prescribed a clopidogrel based regimen (such as clopidogrel monotherapy or a clopidogrel containing duel antiplatelet therapy), and Group 2 which involves patients taking a non-clopidogrel based regimen (such as aspirin or aspirin + low-dose rivaroxaban). Each group will be further subdivided into *CYP2C19* LoF allele carriers and *CYP2C19* LoF allele non-carriers (Table 1).

**Table 1:** Analysis Groups within the PROSPER Study. Estimated frequencies (%) in each group based on historical data are provided.

|  | *CYP2C19* LoF allele carrier<br>(26.2%) | *CYP2C19* LoF allele non-carrier<br>(73.8%) |
| --- | --- | --- |
| 1. Clopidogrel containing anti-platelet therapy (75%) | **A** | **B** |
| 2. Non-clopidogrel containing anti-platelet therapy (25%) | **C** | **D** |

The data will be explored using descriptive statistics as follows:

1. Comparison of baseline characteristics, treatment group and median follow-up time by carrier status for all participants.
2. Flow chart of recruitment to include numbers who withdraw consent and/or missing data by carrier status and treatment group.

The hazard ratio (HR) or incidence rate ratio (IRR) with associated 95% CIs of primary and secondary outcomes will be estimated to compare carrier status (i.e. those who carry and do not carry *CYP2C19* LoF alleles). Statistical models for primary outcomes will be adjusted for *CYP2C19* gene variant carrier status, treatment group (i.e. clopidogrel or non-clopidogrel), age, gender, ethnicity, diabetes, renal disease, smoking status, limb Rutherford classification and prior intervention to the index leg. Subjects will be followed until the primary endpoint or death for any cause with censoring at 12 months. Potential competing events are defined as death from any cause and a sensitivity analysis will be included to assess competing risks for amputation-free survival at 1-year. A significance level of 5% will be used throughout.

Primary and secondary outcomes will be explored as follows:

1. The number of primary and secondary events by carrier status with crude incidence rates of events (with 95% CIs) at 1-year.
2. Kaplan-Meier (KM) plots for both primary outcomes by carrier status.
3. Cumulative Incidence Functions (CIF) and 1-KM function plots for amputation-free survival by carrier status will be compared to investigate potential competing risks^12^.

### Primary outcomes analysis

1. <u>Amputation-free survival at 1-year</u> will be investigated using Cox regression to compare carrier status. The assumption of proportional hazards will be assessed examining Schoenfeld residuals and by including an interaction between time and carrier status in the model^13^. Proportional Hazards (PH) will also be assessed using Mantel-Haenszel methods to compare hazard ratios with the Cox model. If the PH assumption cannot be satisfied, amputation-free survival at 1-year will be analysed using Restricted Mean Survival Time(RMST)^14^. If the comparison for CIF and 1-KM functions suggests the potential for competing risks, then the analysis will examine the cause-specific hazard model, presenting results for both cause-specific HRs and sub-distribution HRs^12^. Both Cox regression and RMST can produce estimates of the HR in the presence of competing risks.
2. <u>Composite of MALE in the index limb</u> [amputation above the ankle, major limb re-intervention (graft revision, thrombectomy or thrombolysis)] <u>and death from any cause at 1-year</u> will be investigated using the same approach as amputation-free survival (excluding the competing risks analysis).

### Secondary outcomes analysis

1. MACE events at any time (MI, CVA or all-cause death) at 1-year will be analysed in the same manner as amputation-free survival (excluding the competing risks analysis).
2. Death from any cause within 30 days. Numbers will be presented by carrier status and treatment group. No formal comparison between carrier status will be made.
3. Minor re-intervention (angioplasty, stent). A zero-inflated negative binomial model will be used to examine the number of minor re-interventions at 1-year. If the alpha parameter and likelihood ratio test from this model demonstrate there is no issue with overdispersion, then minor re-interventions at 1-year will be examined using a zero-inflated Poisson model.
4. Total re-interventions will use the same approach as minor re-interventions at 1-year.
5. Rate of systemic or gastrointestinal bleed will use the same approach as minor re-interventions at 1-year.

### Additional Analyses

1. Exploratory analysis for the interaction of carrier status and treatment group for both primary outcomes. The model chosen will be based on the primary analysis depending on the PH assumption and if competing risks are present for amputation-free survival at 1-year (i.e. either Cox or RMST).
2. If the level of missing data is much higher than expected, multiple imputation using chained regression equations will be used as a sensitivity analysis to compare HRs with the primary analysis.

### Data statement

An anonymised version of the genetic dataset will be stored in a data repository with no limitations on access or use. Genotype data will be deposited in the Figshare repository (https://figshare.com/) at the end of the study following publication. Data will be open access and will be entirely anonymised, with no study ID.

### Patient and public involvement

During protocol development, the study design and methodology were discussed with the patient and public involvement and engagement with research team, called Vocal, at MFT. This was done in a focus group session. The results of the study will be provided back to the Vocal group. Patients participating in the study are provided with an option on the consent form to receive a summary of the study results.

### Ethics and dissemination

Manchester University Research Ethics Committee approval as obtained was part of the IPTIP trial process (IRAS 305751). The results of the study will be published in a peer-review journal and presented at international conferences.

